# Estimating the effectiveness of routine asymptomatic PCR testing at different frequencies for the detection of SARS-CoV-2 infections

**DOI:** 10.1101/2020.11.24.20229948

**Authors:** Joel Hellewell, Timothy W. Russell, The SAFER Investigators and Field Study Team, The Crick COVID-19 Consortium, CMMID COVID-19 working group, Rupert Beale, Gavin Kelly, Catherine Houlihan, Eleni Nastouli, Adam J. Kucharski

## Abstract

**Background:** Routine asymptomatic testing using RT-PCR of people who interact with vulnerable populations, such as medical staff in hospitals or care workers in care homes, has been employed to help prevent outbreaks among vulnerable populations. Although the peak sensitivity of RT-PCR can be high, the probability of detecting an infection will vary throughout the course of an infection. The effectiveness of routine asymptomatic testing will therefore depend on testing frequency and how PCR detection varies over time.

**Methods:** We fitted a Bayesian statistical model to a dataset of twice weekly PCR tests of UK healthcare workers performed by self-administered nasopharyngeal swab, regardless of symptoms. We jointly estimated times of infection and the probability of a positive PCR test over time following infection, we then compared asymptomatic testing strategies by calculating the probability that a symptomatic infection is detected before symptom onset and the probability that an asymptomatic infection is detected within 7 days of infection.

**Findings:** We estimated that the probability that the PCR test detected infection peaked at 77% (54 - 88%) 4 days after infection, decreasing to 50% (38 - 65%) by 10 days after infection. Our results suggest a substantially higher probability of detecting infections 1–3 days after infection than previously published estimates. We estimated that testing every other day would detect 57% (33-76%) of symptomatic cases prior to onset and 94% (75-99%) of asymptomatic cases within 7 days if test results were returned within a day.

**Interpretation:** Our results suggest that routine asymptomatic testing can enable detection of a high proportion of infected individuals early in their infection, provided that the testing is frequent and the time from testing to notification of results is sufficiently fast.

**Funding:** Wellcome Trust, National Institute for Health Research (NIHR) Health Protection Research Unit, Medical Research Council (UKRI)

## Introduction

Detection of current infection with Severe Acute Respiratory Syndrome coronavirus 2 (SARS-CoV-2) is a crucial component of targeted policy responses to the COVID-19 pandemic that involve minimising infection within vulnerable groups. For instance, residents and staff in care homes may be tested regularly to minimise outbreaks among elderly populations^1^. Alternatively, healthcare workers (HCWs) may be routinely tested to prevent nosocomial transmission to patients who may have other comorbidities^2,3^. Both of these populations have a substantially higher risk of fatality from COVID-19 infection than the general population^4,5^.

In the UK, testing commonly uses polymerase chain reaction (PCR) to detect the presence of viral RNA in the nasopharynx of those sampled^6^. The sensitivity of PCR tests at any given point during infection depends upon the amount of viral RNA present, this increases at the start of the infection up to the peak viral load, which appears to occur just before, or at, the time of symptom onset^7–9^. Viral load then decreases, but infected individuals continue to shed the virus for an average of 17 days after initial infection (but this can be far longer than the average, the longest observed duration has been 83 days)^10^. A greater severity of illness is frequently associated with a significantly longer duration of viral shedding^11–13^. Asymptomatic infections have been found to have similar viral loads to symptomatic cases around the time of infection, but instead exhibit shorter durations of viral shedding^14^.

Estimates of temporal variation in the probability of detecting infections by PCR are crucial for planning effective routine asymptomatic testing strategies in settings with vulnerable populations. The testing frequency required to detect the majority of infections before they can transmit onwards will depend on both how soon - and how long - an individual remains positive by PCR test. Measuring the probability that testing will detect SARS-CoV-2 at a given time-since-infection is challenging for two main reasons. First, it requires knowledge of the timing of infection, which is almost always unobserved. Second, it requires a representative sample of tests done on people with and without symptoms performed at many different times with regards to the time of infection. Testing is usually performed on symptomatic infections after symptom onset, leading to an unrepresentative sample^15^.

To address these challenges, we analysed data that covered the regular testing of healthcare workers (HCWs) in London, United Kingdom. We inferred their likely time of infection and used the results of the repeated tests performed over the course of their infection to infer the probability of testing positive depending on the amount of time elapsed since infection. This overcame the bias towards testing around the time of symptom onset, although we focused on data from symptomatic infections so that the timing of symptom onset could be used to infer the likely time of infection.

## Methods

We used data from the SAFER study^16^ conducted at University College London Hospitals between 26 March and 5 May 2020, which repeatedly tested 200 patient-facing HCWs by PCR and collected data on COVID-19 symptoms at the time of sampling^16^. Samples were tested utilising the pipeline established by the Covid-Crick-Consortium. Individuals were asymptomatic at enrollment and were tested for SARS-CoV-2 antibodies at the beginning and end of the study period. Out of the 200 HCWs enrolled in the study, 46 were seropositive at the first antibody test, 36 seroconverted over the study period, and 42 returned a positive PCR test at some point during the study (a detailed analysis of the characteristics of this HCW cohort can be found in Houlihan et al. (2020)). We focused on a subset of 27 of these HCWs that seroconverted during the study period and reported COVID-19 symptoms at one or more sampling times (Figure 1). Combining data on 241 PCR tests performed on self-administered nasopharyngeal samples from these 27 individuals, we estimated the time of infection for each HCW as well as simultaneously estimating the probability of a positive test depending on the time since infection.

**Figure 1:**
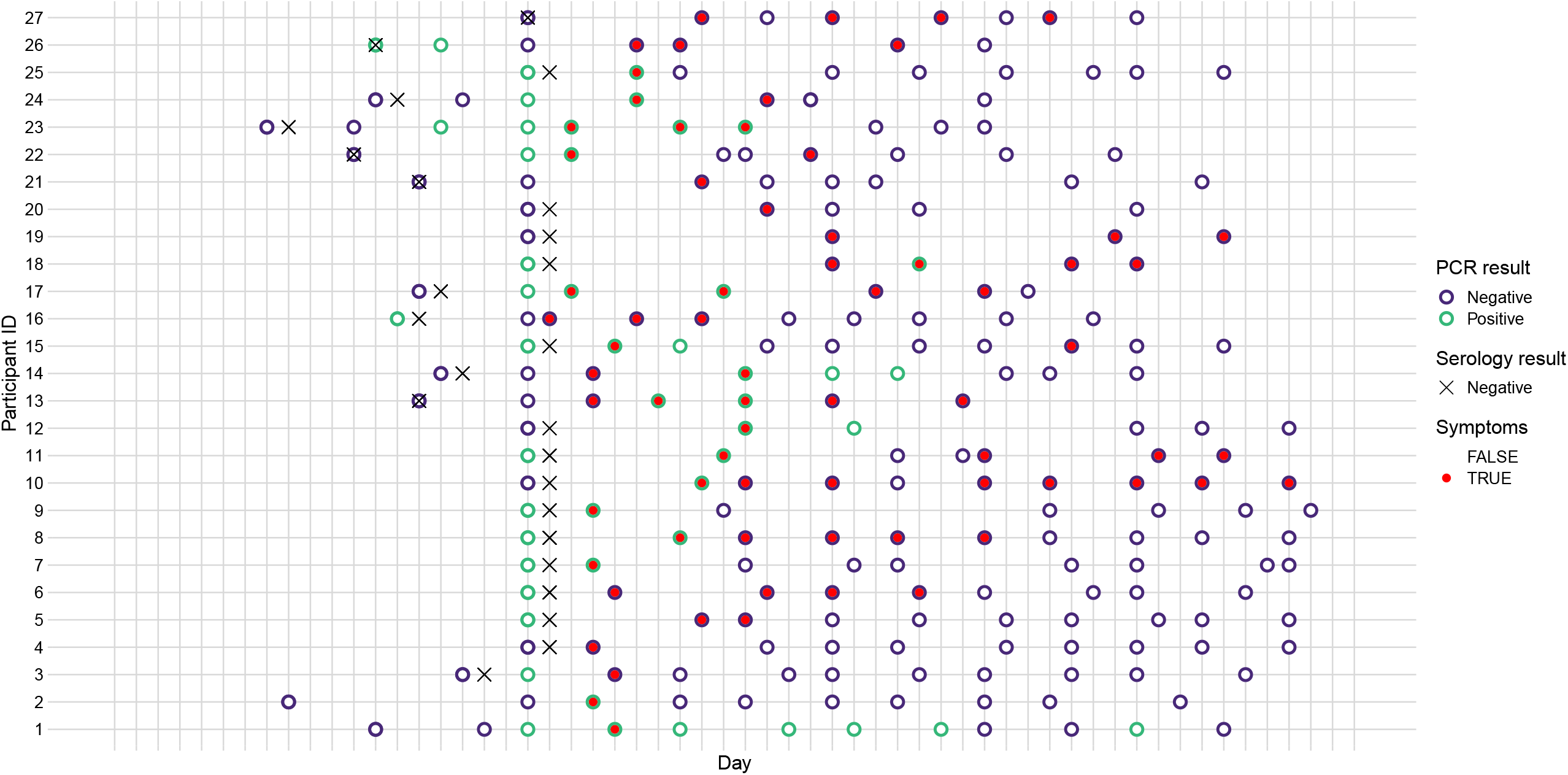
Testing and symptom data for the 27 individuals used in the analysis. Each point represents a symptom report and PCR test result. Red points indicate a positive PCR result while black points indicate a negative PCR result. If any symptoms were reported, the point is triangular while if no symptoms were reported the point is circular. Green crosses show the date of the initial negative serological test. Points are aligned along the x-axis by the timing of each participant’s last asymptomatic report.

We developed a Bayesian model to jointly infer both the likely infection time for each individual and the probability of a positive PCR test depending on the time since infection across all individuals. We used a likelihood function specifically for inferring parameters from censored data^17^ to derive a posterior distribution for the time of infection. This accounts for the fact that the true onset time is censored, i.e. symptom onset for each individual could have occurred anywhere between their last asymptomatic report and their first symptomatic report. Specifically, individual *i* has their likely infection time,*T*_*i*_, inferred based on the interval between their last asymptomatic report, 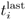, and their first symptomatic report, 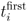. The log-likelihood for the infection time for person *i* is as follows:

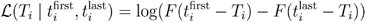

where *F* is the cumulative density function of the lognormal distribution for the incubation period of COVID-19 as estimated in Lauer et al. (2020)^18^. For a detailed description of the procedure used to arrive at the onset times from the censored data and list of the sources of uncertainty in our model, see Supplement D.

For a given inferred infection time for person *i*, the relationship between the time since infection and a positive PCR test on person 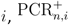, administered at time *t*_*n,i*_ is given by a piecewise logistic regression model with a single breakpoint:

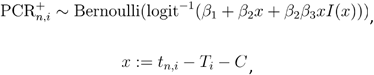

Where *C* is the time of the breakpoint, *x* is the amount of time between infection and testing minus the value of the breakpoint, *I(x)* is a step function that equals 0 if or equals *x <* 1 if, and the *β* terms define the regression coefficients fit across all tests and people.

To ensure biological plausibility, each individual was assumed to have a negative result at their precise time of infection to constrain the PCR positivity curve to have 0 probability of detection at 0 days since infection. We fitted the model using R 4.0.3^19^ and Stan 2.21.2^20^, the data and the code required to reproduce the figures and results of this study can be found at the public github repository: https://github.com/cmmid/pcr-profile. We ran four MCMC chains for 2000 samples each, discarding the first 1000 samples from each chain as warm-up iterations. Convergence of the chains was assessed using the R-hat statistic being 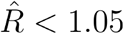 for each model parameter.

We also performed a sensitivity analysis whereby the testing data for one HCW at a time was left out from the model fitting procedure to see if the PCR testing data for any individual HCW had an undue influence on the overall regression fit (results are shown in Supplement B).

We looked at two different ways of assessing the performance of different routine asymptomatic testing frequencies. Firstly, we calculated the probability that a symptomatic case would be detected before symptom onset; this demonstrates the ability of testing to catch infections before people eventually self-isolate due to symptoms (by which point they may already have infected someone). Secondly, we calculated the probability that an asymptomatic case is caught within 7 days of infection, estimating how frequently testing would need to be to detect asymptomatic infections in a timely manner. The mathematical equations used to calculate each of these probabilities are shown in Supplement C.

## Results

The model found that the majority of individuals included in this analysis were infected around the beginning of the study period in late March (Figure 2). This corresponds with a period of greatly increased hospitalisation in London, which could potentially mean much higher exposure to infectious COVID-19 patients. However, this analysis cannot say for certain where these HCWs were infected.

**Figure 2:**
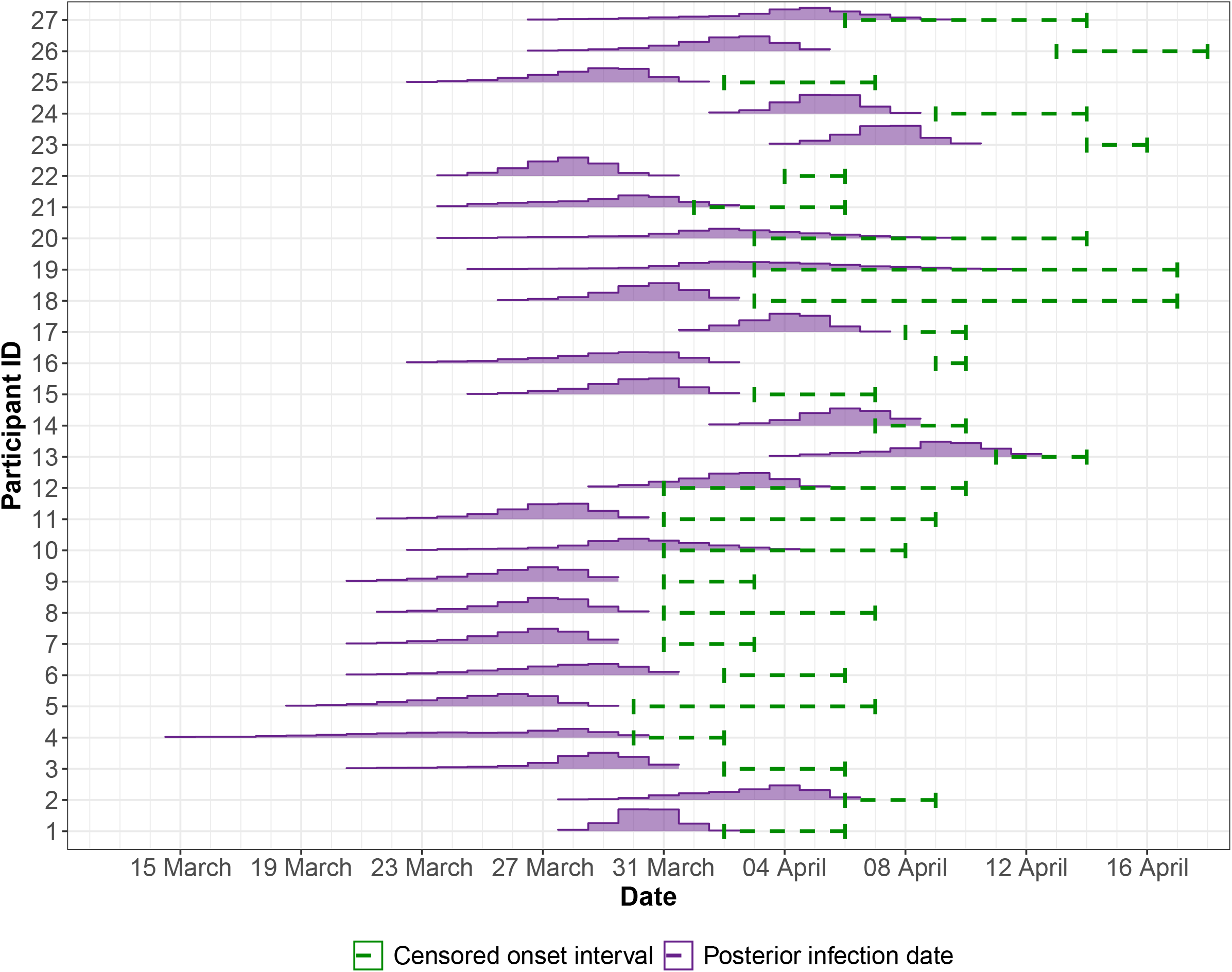
The posterior of the infection time (Ti) of each participant. The posterior distribution of the infection time for each participant (purple) alongside the censored interval within which their symptom onset occurred (green dashed lines).

We estimated that the peak probability of a positive PCR test is 77% (54 - 88%) at 4 days after infection. The probability of a positive PCR test then decreases to 50% (38 - 65%) by 10 days after infection and reaches virtually 0% probability by 30 days after infection (Figure 3B). Summary statistics for the posterior distributions of the piecewise logistic regression parameters are shown in Table 1. We compared our results for the probability of infection throughout infection to previous results in Supplement A, we found greater probability of detecting infections 1 to 3 days infection and a consistently lower probability of detection infections around 10 to 30 days after infection than previous results.

**Table 1:** Summary of model parameters and the median and 95% credible interval from their fitted posterior distributions.

| Parameter | Description | Interpretation | Posterior median (95% credible interval) |
| --- | --- | --- | --- |
| $C$ | Breakpoint of piecewise regression | The time at which PCR positivity begins to peak | 3.18 days post-infection (2.01 - 5.11) |
| $\beta_1$ | Intercept of both regression curves | N/A | 1.51 (0.80— 2.31) |
| $\beta_2$ | Slope of 1st regression curve | The rate of increase in percentage of infections detected after exposure | 2.19 (1.26—3.47) |
| $\beta_3$ | Slope of 2nd regression curve | The rate of decrease in the percentage of infections detected, after the curve peaks | -1.1 (-1.2—-1.05) |

**Figure 3:**
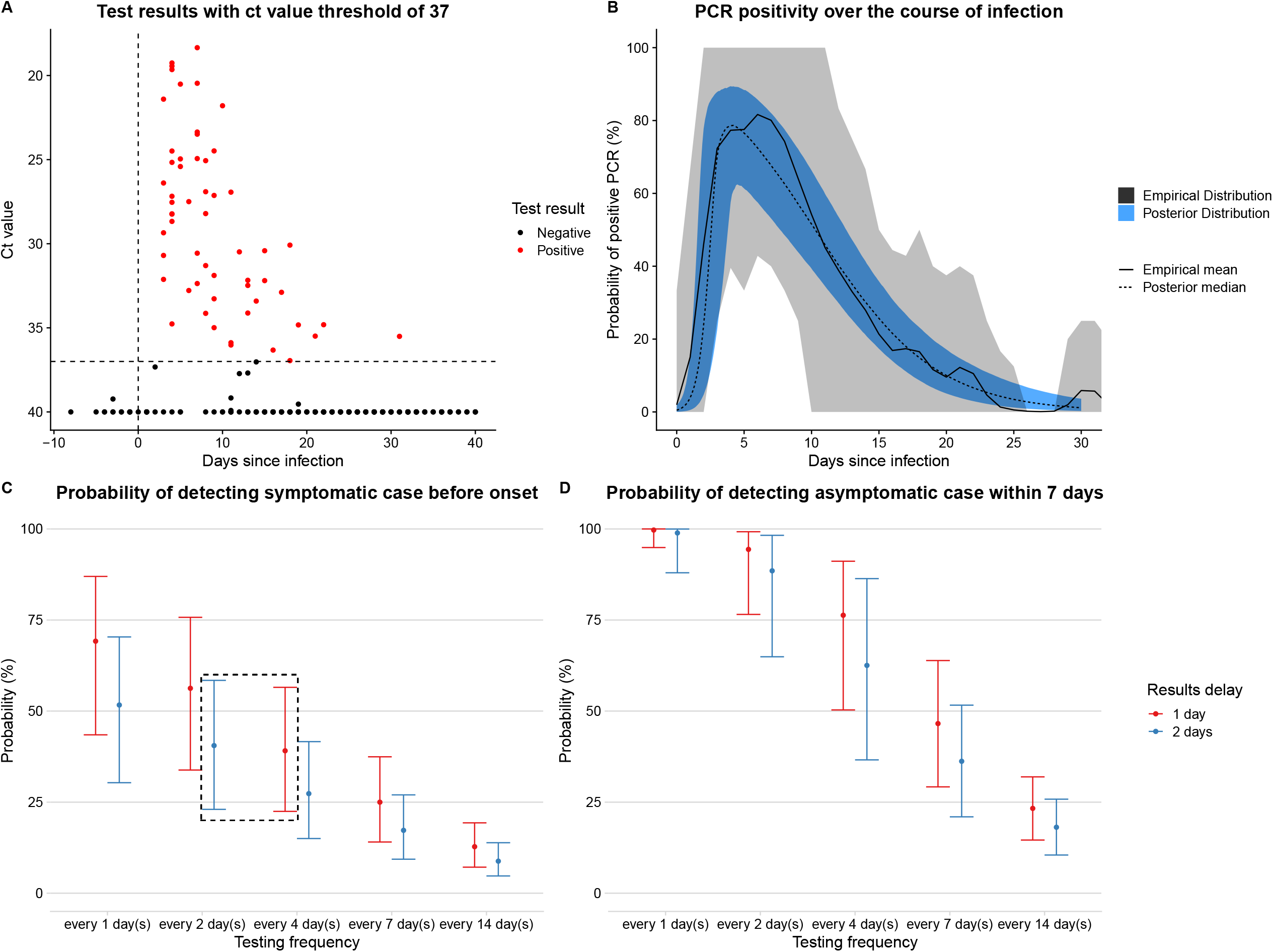
Estimation of positivity over time, and probability that different testing frequencies with PCR would detect virus. A) Ct value data for the PCR tests in the SAFER trial. This plot does not show data for every individual included in the analysis. The x-axis shows a time since infection using the median infection date inferred by the model. Points above the threshold of 37, indicating a positive result, are shown in red. Negative results above 37 are shown in black. All negative results for which there is no ct value specified are given the value of 40. B) Temporal variation in PCR-positivity based on time since infection. The grey interval and solid black line show the 95% uncertainty interval and the mean, respectively, for the empirical distribution calculated from the posterior samples of the times of infection (see Supplement D1 for methodology). The blue interval and dashed black line show the 95% credible interval and median, respectively, of the logistic piecewise regression described above. C) Probability of detecting virus before expected onset of symptoms, based on curve in (B), assuming delay from test to results is either 1 or 2 days. Dashed black box shows a site of possible trade-off between testing frequency and results delay discussed in the text D) Probability of detecting an asymptomatic case within 7 days, based on curve in (B), assuming delay from test to results is either 24 or 48 hours.

Our routine asymptomatic testing scenarios established that the higher the frequency of testing, the higher the probability that a symptomatic case will be detected before symptom onset (Figure 3C) and the higher the probability that an asymptomatic case is detected within 7 days (Figure 3D). A 2 day delay between testing and notification compared to a 1 day delay led to reduced probability of detection in both testing scenarios (Figures 3C, 3D). This is because a longer delay means that an infection must be caught earlier to allow for a longer period of time between a test being administered and the infected person being notified of the results. An increased delay from testing to notification caused a greater relative reduction in the probability of detecting an asymptomatic case within 7 days of infection when the testing frequency was lower (Figure 3D).

When considering what is an acceptable testing frequency for detecting a desired proportion of symptomatic cases prior to their symptom onset, there may be a trade-off between testing frequency and the delay from testing to notification. For example, the probability of detecting a symptomatic case prior to onset is very similar for a 2 day testing frequency with a 2 day notification delay (41%, 23 - 58%) compared to a 4 day testing frequency with a 1 day notification delay (39%, 22 - 56%). This trade-off is depicted graphically in the dashed black box in Figure 3B.

During 2020, lateral flow tests (LFTs) with a turnaround time of roughly 30 minutes for the detection of SARS-CoV-2 have been developed and evaluated^21^. Such tests typically have a lower mean sensitivity than standard PCR tests. However, the faster turnaround time can aid the logistical challenge posed by rapid large-scale testing. Thus far in our analysis, a positive PCR test has been defined by a cycle threshold (Ct) value of less than or equal to 37. However, given that Ct values are also available for the tests in our dataset, we were able to redefine test outcomes using different Ct value thresholds that reflect the potential sensitivity of the more recent LFTs, which can generally detect infectiousness (when viral loads are high) but not always infection (when viral loads may be lower)^22^.

The model was re-fitted using two potential LFT-like definitions of a positive test: a Ct value of less than or equal 28, or less than or equal to 25. The newly defined test outcomes are shown in panel A of Figures 3-S1 & 3-S2, along with the corresponding estimates of test sensitivity as a function of time since infection in panel B. We then used the sensitivity curves in the symptomatic and asymptomatic testing scenarios with frequent testing, assuming no delay between rapid test and result (reflecting the imagined use case of LFTs, results shown in panels C and D).

For the hypothetical LFT test scenario compared to the PCR tests, the peak probability of detection is lower, with a peak probability of detection of 64% (33 - 85%) at 4.3 days after infection and 42% (13 - 70%) at 3.8 days after infection for Ct values of 28 and 25, respectively. The probability of detection by LFT also declines to negligible values far sooner after infection, by around 18 days, compared to around 30 days for PCR. However, the uncertainty in the probability of detection curve is wider for these hypothetical LFT tests compared to PCR because there were fewer positive tests to fit to overall. The probability of detecting symptomatic cases before symptom onset, or asymptomatic cases within 7 days of infection, decreases when the Ct threshold for a positive test is lower (panels C and D of Figures 3-S1 and 3-S2). When the Ct threshold is defined to be 25, even testing every two days yields a median probability of detecting symptomatic cases before onset below 50%.

## Discussion

The ongoing COVID-19 pandemic has led to increasing focus on routine asymptomatic testing strategies that could prevent sustained transmission in hospitals and other defined settings with at-risk individuals such as care homes. Using data on repeated testing of healthcare workers, we estimated that peak positivity for PCR tests for SARS-CoV-2 infections occurs 4 days after infection, which is just before the average incubation duration, in agreement with other studies finding that viral load in the respiratory tract is highest at this point^23,24^.

We found a substantially higher probability of detection by PCR between 1 and 3 days after infection than a previous study^25^. The low detection probabilities estimated in the previous study for the period 1 to 3 days after infection were fitted to very small amounts of data: one observed negative test on each of 1, 2, and 3 days after infection. Due to the fact that HCWs in the SAFER study were repeatedly tested even when asymptomatic, many of the tests took place close to the inferred infection times. This provided more test data for our model to fit to for the period just after infection. We provide a more rigorous exploration of the differences between our results and existing work in Supplement A.

Our model also estimated much lower probabilities of detection between 7 and 30 days after infection compared to the models by Kucirka et. al. and Hay & Kennedy-Schaffer et. al. A plausible explanation for this difference could be due to the sample collection method and disease severity of the people being tested, leading to different observed viral load dynamics. The SAFER study data used here was collected from self-administered tests by HCWs and the symptoms recorded were those that were compatible with SARS-CoV-2 according to Public Health England, including a “new continuous cough or alteration in sense of taste or smell”^16^. Conversely, the datasets used for fitting the Kucirka model consist mainly of HCW-administered tests on hospitalised patients who are likely to have more severe infections, a factor that has been associated with a longer duration of viral shedding^10^ in some studies. As such, our curve for the probability of detection by PCR may constitute a closer approximation of PCR test sensitivity over time in individuals with mild symptomatic infections. This would make it particularly useful for estimating the effectiveness of routine asymptomatic testing strategies, which would seek to detect all infections, not just the most severe.

Incorporating our estimates of PCR detection probability into a model of routine asymptomatic testing strategies, we found that there is the potential for a trade-off between the turnaround time for test results and testing frequency (Example in dashed black box, Figure 3C). This could be particularly relevant for settings that do not have the resources or capacity for very high frequency testing, but could ensure prompt results. Although our analysis focuses on the probability of testing positive, any potential testing and isolation strategy would also need to consider the potential for false positives, particularly at low prevalence^26^.

The maximum probability of detection of 77% shown by the curve in Figure 3B refers to the whole population and does not imply that an individual person’s peak probability of being detected by a PCR test is 77%. The curve is fitted to combined test results for many individuals, each of whom will have had variation in the timing of their particular peak probability of detection. This variation is smoothed out over all individuals to lead to the curve shown in Figure 3B.

To explore the potential for rapid testing of individuals, we examined how the curve in Figure 3B would change if the cycle threshold used to define a positive result was lowered, which mimics the detection capabilities of lateral flow tests that are less able to detect infections at higher Ct values^22,27^. We estimated that the probability of detection post-infection still peaks around 4 days after infection, but that the peak probability of detection is lower and the probability of detection declines much faster after the peak. The reduced period of time after infection during which a case might be detected in our hypothetical LFT scenario compared to PCR may help to explain some of the low sensitivities for LFTs reported during the evaluation of LFT testing programmes such as in Liverpool, where LFTs detected only 48.89% of the infections that were later confirmed by PCR^28^. In general, our estimates correspond with previous observations that infections with lower viral loads (which are likely to be older infections and will have higher Ct values) are less likely to be detected by LFTs compared to PCR.

We assumed that symptoms reported during the study were due to clinical episodes of COVID-19 infection, and not due to other respiratory infections with similar symptoms. All individuals in the analysis seroconverted over the course of the study, suggesting that such symptoms were likely to be associated with SARS-CoV-2 infection.

Our analysis is also limited by excluding asymptomatic HCWs that seroconverted over the course of the study. Symptomatic infections may have higher viral loads and be more likely to be detected than asymptomatic infections, however this has not been found to be the case elsewhere^14^. Our repeated testing model presents results for detecting asymptomatic infections that relies on the assumption that the probability of detection over time is the same for symptomatic and asymptomatic infections. If asymptomatic infections are instead less likely to be detected then our estimate of the probability of detection within 7 days of infection will be an overestimate.

Routine asymptomatic testing is a crucial component of effective targeted control strategies for COVID-19, and our results suggest that frequent testing and fast turnaround times could yield high probabilities of detecting infections – and hence prevent outbreaks – early in at-risk settings.

## Data Availability

All of the data and the code required to reproduce the figures and results of this study can be found at the public github repository: https://github.com/cmmid/pcr-profile.

https://www.thelancet.com/pdfs/journals/lancet/PIIS0140-6736(20)31484-7.pdf

## Data availability

The subset of the data, including individuals that seroconvert and show symptoms at some stage during the data collection period, required to reproduce the figures and results of this study can be found at the public github repository: https://github.com/cmmid/pcr-profile.

## Code availability

The code required to reproduce the figures and results of this study can be found at the public github repository: https://github.com/cmmid/pcr-profile.

## Acknowledgments

The following funding sources are acknowledged as providing funding for the named authors. Wellcome Trust (206250/Z/17/Z: AJK, TWR; 210758/Z/18/Z: JH). AK was supported by the NIHR HPRU in Modelling and Health Economics, a partnership between PHE, Imperial College London and LSHTM (grant code NIHR200908). The views expressed are those of the authors and not necessarily those of the United Kingdom (UK) Department of Health and Social Care, the National Health Service, the National Institute for Health Research (NIHR), or Public Health England (PHE). The SAFER study was funded by MRC UKRI (grant MC_PC_19082) and supported by the UCLH/UCL NIHR BRC.

---

**Figure 3-S1/S2:**
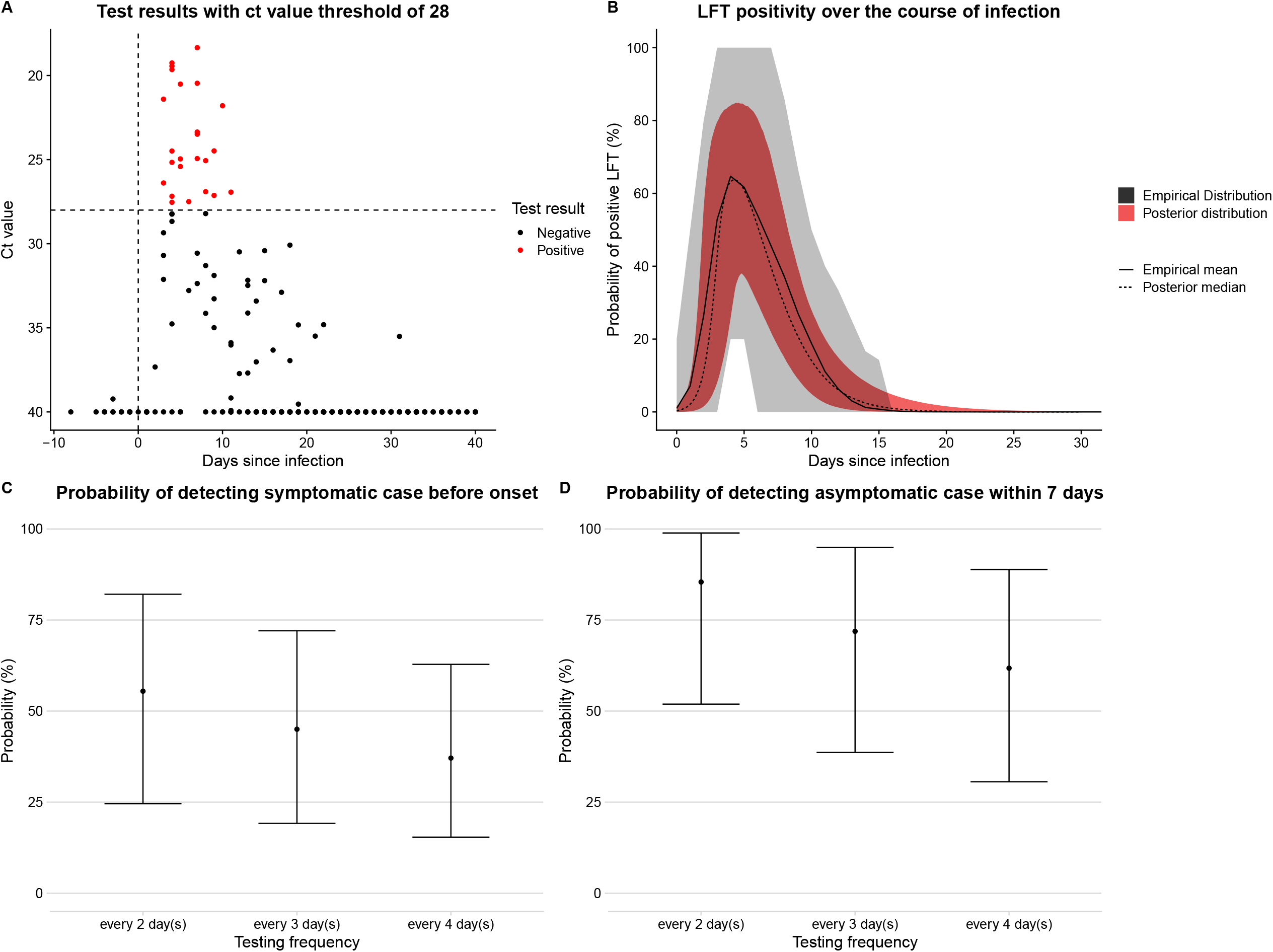
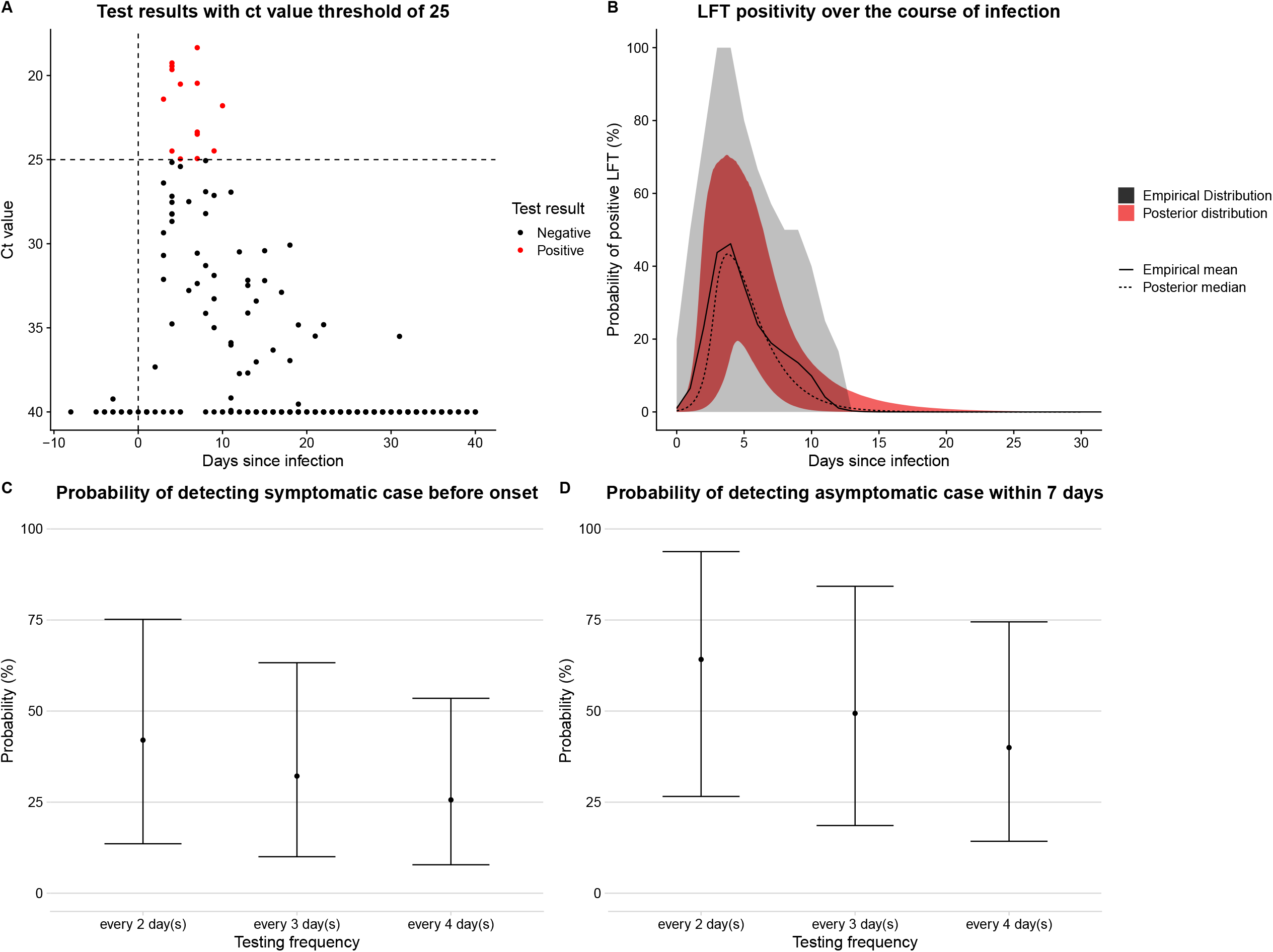
A copy of Figure 3 using a Ct value of 28 or 25 (instead of 37) to classify a test as positive or not. This is instructive of how a lateral flow test (LFT) might perform as they seem to be less sensitive to infections with lower viral loads than PCR tests. In panels C and D the probabilities of detection are now considered with a 0 day delay since LFTs give results within minutes that can be passed on to the person being tested quickly.

