## Supplementary material for "Estimating the effectiveness of routine asymptomatic PCR testing at different frequencies for the detection of SARS-CoV-2 infections"

Members of all three consortia are listed in Supplements E, F, and G.

Corresponding author: Joel Hellewell, Faculty of Epidemiology and Population Health, London School of Hygiene & Tropical Medicine, Keppel Street, London WC1E 7HT, United Kingdom..

### A. Comparison with previous results

We compared our findings for the probability of detection by PCR as a function of time since infection to two existing[1,2] results (Figure S1A). Kucirka et al. fit a polynomial curve to results from 8 studies that estimated PCR sensitivity as a function of time since symptom onset or exposure. Hay & Kennedy-Schaffer fit a model motivated by prior knowledge of viral load dynamics to PCR sensitivity as a function of symptom onset as compiled by Borremans et al.[3]

Our model, fitted to the SAFER study data, found a much higher probability of detecting infections between 1 and 3 days after infection than the model fitted by Kucirka et. al. The peak probability of detection was also later on during the course of infection for Kucirka et. al, who estimated it to be around 8 days after infection compared to 4 days found by our model and Hay & Kennedy-Schaffer et. al. Our estimated probability of detection was also consistently lower than that found by both other models for 7 days after infection onwards.

We summarised the SAFER study data using the median infection times inferred by our model to provide point estimates of the observed probability of detection for each day since infection (shown in the orange points in Figure S1A). We then re-fit the Kucirka model to its original data set extended to include the HCW data from the SAFER study (Figure S1B). Doing this gave estimates of the probability of detection between 1 and 3 days after infection from the Kucirka model that were very similar to those found by our model.

However, the Kucirka model still estimates higher probabilities of detection than our model from 7 days after infection onwards. This seems to be a feature of the SAFER study data rather than a poor model fit to the SAFER data (Figure S1A).

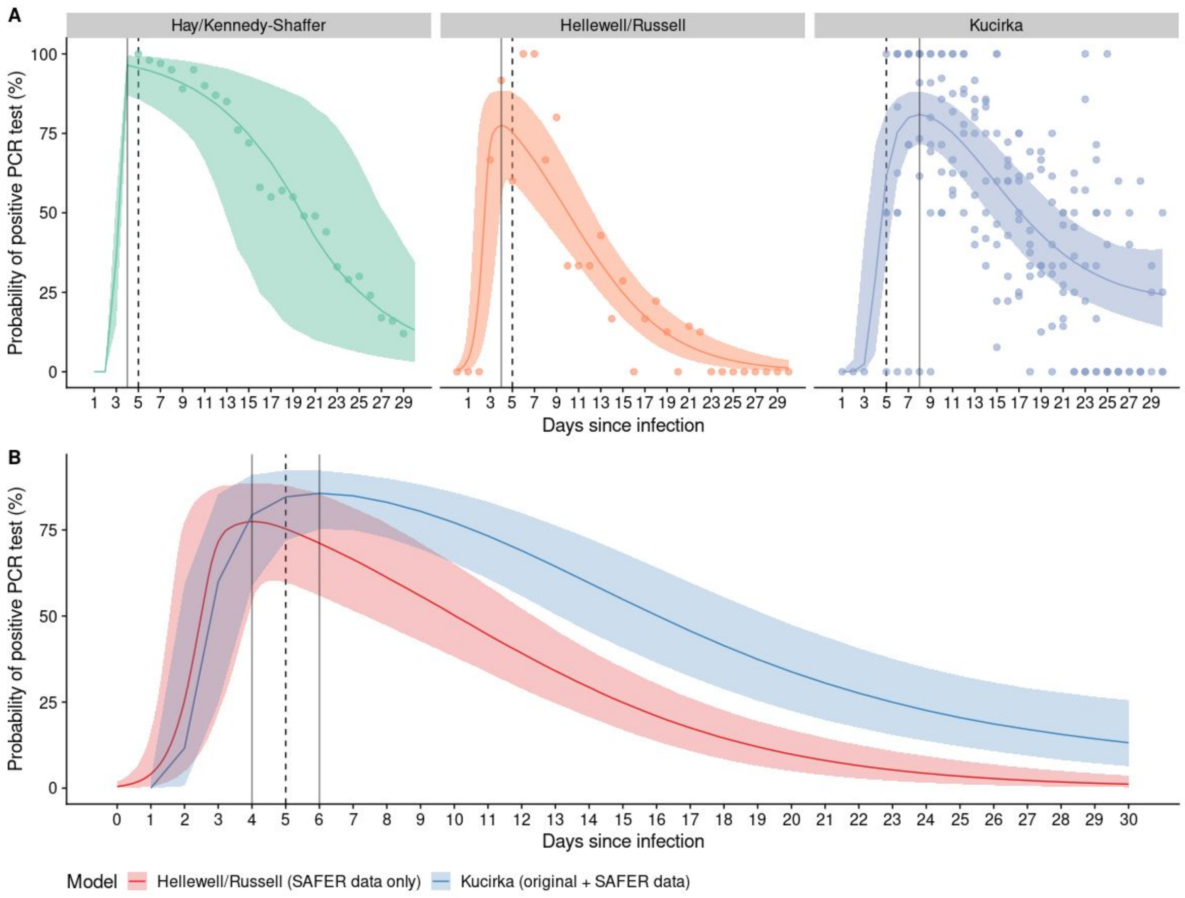


**Figure S1:** A) Three PCR detection probability curves as a function of the time since infection along with data used to fit the curve. From L - R: (Green) A curve fit to data from Borremans et al (2020) by Hay & Kennedy-Shaffer et al (2020). (Orange) Our curve fit to data on HCWs, the points show the estimated sensitivity of PCR tests at each time point using the median infection time estimated by our model. (Purple) The curve fit Kucirka et al (2020) to data combined from 8 studies. B) A copy of our PCR detection probability curve (same as orange curve in A) is shown in red. The blue curve shows the results of fitting the model developed by Kucirka et al 2020 to their data (purple points in A) combined with our data (orange points in A).

### B. Sensitivity Analysis

It is possible that the fitting procedure that led to our PCR positivity curve over time was heavily influenced by a single individual, given that our curve was jointly fit to the posterior distributions of all individuals (representing the likely time at which they were exposed to SARS-CoV-2). Therefore, to check the influence of each individual on the resulting curve, we performed a leave-one-out sensitivity analysis, whereby we fit the same curve to the 27 possible sets of 26 individuals if one individual is left out in each fit.

The medians of the resulting 27 fits and their corresponding 95% credible intervals are plotted in Figure S2, where it is possible to see that each run is largely in agreement with all of the others. In particular, the 95% credible intervals of all curves are in almost exact agreement. The median of one curve is noticeably lower than the others after the 11-12 day mark. However, on inspection of the 95% credible interval for this curve in comparison to the other 26, it is possible to see that this variation is captured within the 95% credible intervals of the other curves. Specifically, we notice that the 95% credible interval of the curve with a lower median is almost identical to the 95% credible intervals of the other 26 curves.

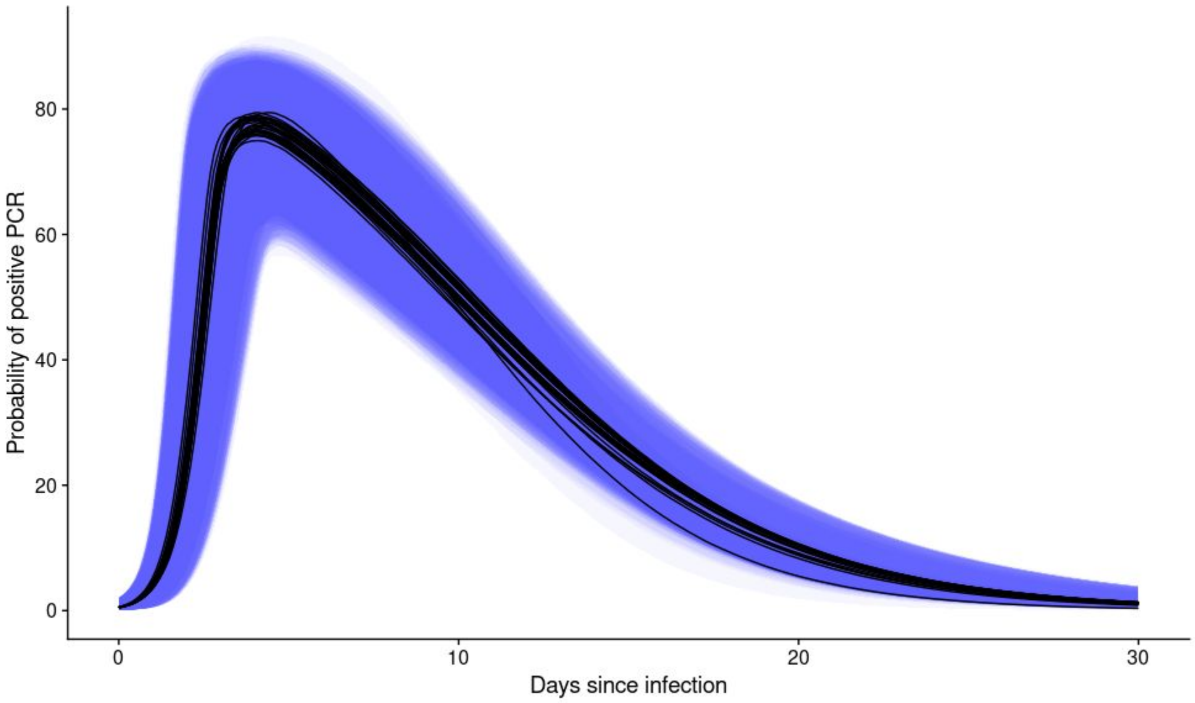


**Figure S2: Multiple PCR positivity curves superimposed on top of each other, each curve shows the fitted PCR positivity curve while leaving out data for a different one of the 27 individuals in the data set each time.** There is one curve whereby the median posterior probability is around 5% lower from ~12 days after infection onwards if data for an individual is excluded. This suggests that one individual out of the 27 HCWs continued to test positive for a long time after their inferred infection date, which could possibly bias our PCR positivity upwards slightly towards the tail of the distribution.

### C. Routine Asymptomatic Testing Model

To calculate the probability that a symptomatic infection is detected prior to symptom onset, let $I$ be the set of the possible testing times for a given test frequency $f_x$, which given explicitly, can be written as

$$I = \{[0, f_x, 2f_x, \ldots], [1, f_x+1, 2f_x+1, \ldots], \ldots, [f_x-1, 2f_x-1, 3f_x-1 \ldots]\}$$

The maximum values of $i \in I$ are set at 30 since testing PCR positive 30 days after infection is unlikely.

For the given testing times $i \in I$, if we denote the $j_{th}$ testing time in $i$ as $i_j$, the number of testing times in $i$ as $|i|$, and $d$ as the delay between test and result, the probability of detecting an infection before symptom onset for testing times $i$ is equal to

$$P_i = \sum_{j=1}^{|i|} g(i_j)p(i_j - d) \prod_{k=j-1}^{0} \Big( (1 - g(i_k))(1 - p(i_k - d)) \Big)$$

where $g(t)$ is the probability of no onset before time $t$ and $p(t)$ is the probability of a positive test at time $t$.

Noting that $|I| = f_x$, the probability of detecting a symptomatic infection before symptom onset over all possible testing time variations $i \in I$ is therefore

$$\frac{1}{f_x} \sum_{i \in I} P_i$$

For asymptomatic infections, the value of $g(t) = 1 \ \forall t$ because there will never be an onset time. For detection within seven days we consider

$$I^* = \{[0, f_x, 2f_x, \ldots], [1, f_x+1, 2f_x+1, \ldots], \ldots, [f_x-1, 2f_x-1, 3f_x-1 \ldots]\}$$

with values up to $7 - d$, since a positive test needs to be performed by this point to be returned within 7 days. For the given testing times $i \in I^*$, the probability of detecting an asymptomatic infection within 7 days is

$$P_i^* = \sum_{j=1}^{|i|} p(i_j) \prod_{k=j-1}^{0} 1 - p(i_k) \quad ,$$

and lastly the probability of detecting an asymptomatic infection within 7 days over all testing time variations $i \in I^*$ is equal to

$$\frac{1}{f_x} \sum_{i \in I^*} P_i^* \quad .$$

### D. Further Methodology

#### 1. Empirical distribution of PCR positivity

The grey interval in Figure 3A is calculated from the posterior samples of the likely infection time for each individual ($T_i$). If we let the $j_{th}$ posterior sample of $T_i$ be denoted $T_{ij}$, then $d_{ij}$, which denotes the time from infection until each test $t_{n,i}$, performed on individual $i$, for each sample $T_{ij}$ is given by

$$d_{ij} = t_{n,i} - T_{ij} \, .$$

Each $d_{ij}$ is rounded to the nearest discrete day and for each MCMC iteration, $j$¸ we calculate the proportion of tests where $d_{ij} = k$ which were positive for each discrete day $k$ since infection, denoted $p_j(k)$. We then calculate the mean and 95% uncertainty intervals of $p_j(k)$ for each day $k$ over all MCMC samples $j$. This can be considered a graphical representation of the "data" that the PCR positivity regression is fit to (the precise values rely on the infection time draws at each iteration of the MCMC).

#### 2. Sources of uncertainty

We infer likely infection times using a Bayesian inference framework, where we are able to include sources of uncertainty in a statistically robust manner. Such sources of uncertainty include:

- The censored nature of the interval between the *last asymptomatic report* and *first symptomatic report* for all individuals
- The incubation period, which is assumed to be a Gamma distribution parameterised using fitted estimates from Lauer et al. (2020)[5]
- As the data to which we fit the curve to is sampled from the inferred posterior distributions corresponding to the likely time at which individuals were infected, the 95% credible interval of our PCR positivity curve over time includes the uncertainty within each of the posterior distributions of likely infection time

### E. The SAFER Investigators and Field Study Team

Rebecca Matthews, Abigail Severn, Sajida Adam, Louise Enfield, Angela McBride, Kathleen Gärtner, Sarah Edwards, Fabiana Lorencatto, Susan Michie, Ed Manley, Maryam Shahmanesh, Hinal Lukha, Paulina Prymas, Hazel McBain, Robert Shortman, Leigh Wood, Claudia Davies, Bethany Williams, Emilie Sanchez, Daniel Frampton, Matthew Byott, Stavroula M Paraskevopoulou, Elise Crayton, Carly Meyer, Nina Vora, Triantafylia Gkouleli, Andrea Stoltenberg, Veronica Ranieri, Tom Byrne, Dan Lewer, Andrew Hayward, Richard Gilson, Naomi Walker.

### F. The Crick COVID-19 Consortium

Aaron Ferron, Aaron Sait, Abhinay Ramaprasad, Abigail Perrin, Adam Sateriale, Adrienne E Sullivan, Aileen Nelson, Akshay Madoo, Alana Burrell, Aleksandra Pajak, Alessandra Gaiba, Alice Rossi, Alida Avola, Alison Dibbs, Alison Taylor-Beadling, Alize Proust, Almaz Huseynova, Amar Pabari, Amelia Edwards, Amy Strange, Ana Cardoso, Ana Agua-Doce, Ana Perez Caballero, Anabel Guedan, Anastacio King Spert Teixeira, Anastasia Moraiti, Andreas Wack, Andrew Riddell, Andrew Buckton, Andrew Levett, Andrew Rowan, Angela Rodgers, Ania Kucharska, Anja Schlott, Annachiara Rosa, Annalisa D'Avola, Anne O'Garra, Anthony Gait, Antony Fearns, Beatriz Montaner, Belen Gomez Dominguez, Berta Terré Torras, Beth Hoskins, Bishara Marzook, Bobbi Clayton, Bruno Frederico, Caetano Reis e Sousa, Caitlin Barns-Jenkins, Carlos M Minutti, Caroline Oedekoven, Catharina Wenman, Catherine Lambe, Catherine Moore, Catherine F Houlihan, Charles Swanton, Chelsea Sawyer, Chloe Roustan, Chris Ekin, Christophe Queval, Christopher Earl, Claire Brooks, Claire Walder, Clare Beesley, Claudio Bussi, Clementina Cobolli Gigli, Clinda Puvirajasinghe, Cristina Naceur-Lombardelli, Dan Fitz, Daniel M Snell, Dara Davison, David Moore, Davide Zecchin, Deborah Hughes, Deborah Jackson, Dhruva Biswas, Dimitrios Evangelopoulos , Dominique Bonnet, Edel C McNamara, Edina Schweighoffer, Effie Taylor, Efthymios Fidanis, Eleni Nastouli, Elizabeth Horton, Ellen Knuepfer, Emine Hatipoglu, Emma Russell, Emma Ashton, Enzo ZPoirier, Erik Sahai, Fatima Sardar, Faye Bowker, Fernanda Teixeira Subtil, Fiona McKay, Fiona Byrne, Fiona Hackett, Fiona Roberts, Francesca Torelli, Ganka Bineva-Todd, Gavin Kelly, Gee Yen Shin, Genevieve Barr, George Kassiotis, Georgina H Cornish, Gita Mistry, Graeme Hewitt, Graham Clark, Gunes Taylor, Hadija Trojer, Harriet B Taylor, Hector Huerga Encabo, Heledd Davies, Helen M Golding, Hon-Wing Liu, Hui Gong, Jacki Goldman, Jacqueline Hoyle, James Fleming, James I MacRae, James M Polke, James M A Turner, JaneHughes, Jean D O'Leary, Jernej Ule, Jerome Nicod, Jessica Olsen, Jessica Diring, Jill A Saunders, Jim Aitken, Jimena Perez-Lloret, Joachim M Matz, Joanna Campbell, Joe Shaw, John Matthews, Johnathan Canton, Joshua Hope, Joshua Wright, Joyita Mukherjee, Judith Heaney, Julian AZagalak, Julie A Marczak, Karen Ambrose, Karen Vousden, Katarzyna Sala, Kayleigh Richardson, Kerol Bartolovic, Kevin W Ng, Konstantinos Kousis, Kylie Montgomery, Laura Churchward, Laura Cubitt, Laura Peces-Barba Castano , Laura Reed, Laura E McCoy, Lauren Wynne, Leigh Jones, Liam Gaul, Lisa Levett, Lotte Carr, Louisa Steel, Louise Busby, Louise Howitt, Louise Kiely,

Lucia Moreira-Teixeira, Lucia Prieto-Godino, Lucy Jenkins, Luiz Carvalho, Luke Nightingale, Luke Wiliams, Lyn Healy, Magali S Perrault, Malgorzata Broncel, Marc Pollitt, Marcel Levi, Margaret Crawford, Margaux Silvestre, Maria Greco, Mariam Jamal-Hanjani, Mariana Grobler, Mariana Silva Dos Santos, Mark Johnson, Mary Wu, Matthew Singer, Matthew J Williams, Maximiliano G Gutierrez, Melanie Turner, Melvyn Yap, Michael Howell, Michael Hubank, Michael J Blackman, Michael D Buck, Michele S Y Tan, Michelle Lappin, Mimmi Martensson, Ming Jiang, Ming-Han C Tsai, Ming-Shih Hwang, Mint RHtun, Miriam Molina-Arcas, Moira J Spyer, Monica Diaz-Romero, Moritz Treeck, Namita Patel, Natalie Chandler, Neil Osborne, Nick Carter, Nicola O'Reilly, Nigel Peat, Nikhil Faulkner, Nikita Komarov, Nisha Bhardwaj, Nnennaya Kanu, Oana Paun, Ok-Ryul Song, Olga O'Neill, Pablo Romero Clavijo, Patrick Davis, Patrick Toolan-Kerr, Paul Nurse, Paul Kotzampaltiris, Paul C Driscoll, Paul R Grant, Paula Ordonez Suarez, Peter Cherepanov, Peter Ratcliffe, Philip Hobson, Philip A Walker, Pierre Santucci, Qu Chen, Rachael Instrell, Rachel Ambler, Rachel Ulferts, Rachel Zillwood, Raffaella Carzaniga, Rajnika Hirani, Rajvee Shah Punatar, Richard Byrne, Robert Goldstone, Robyn Labrum, Ross Hall, Rowenna Roberts, Roy Poh, Rupert Beale, Rupert Faraway, Sally Cottrell, Sam Barrell, Sam Loughlin, Samuel McCall, Samutheswari Balakrishnan, Sandra Segura-Bayona, Savita Nutan, Selvaraju Veeriah, Shahnaz Bibi, Sharon P Vanloo, Simon Butterworth, Simon Caidan, Solene Debaisieux, Sonia Gandhi, Sophia Ward, Sophie Ridewood, Souradeep Basu, Stacey-Ann Lee, Steinar Halldorsson, Stephanie Nofal, Steve Gamblin, Steve Hindmarsh, Stuart Kirk, Subramanian Venkatesan, Sugera Hashim, Susanne Herbst, Suzanne Harris, Svend Kjaer, Tammy Krylova, Tea Toteva, Theo Sanderson, Theresa Higgins, Thomas Martinez, Timothy Budd, Tom Cullup, Venizelos Papayannopoulos, Vicky Dearing, Vijaya Ramachandran, Wai Keong Wong, Wei-Ting Lu, Yiran Wang, Yogen Patel, Zena Collins, Zheng Xiang, Zoe Allen, Zoe H Tautz-Davis.

### G. CMMID COVID-19 Working Group

The following authors were part of the Centre for Mathematical Modelling of Infectious Disease COVID-19 working group. Each contributed in processing, cleaning and interpretation of data, interpreted findings, contributed to the manuscript, and approved the work for publication: Amy Gimma, W John Edmunds, Carl A B Pearson, Kiesha Prem, James D Munday, Katharine Sherratt, Naomi R Waterlow, Graham Medley, Billy J Quilty, Kaja Abbas, Akira Endo, Kathleen O'Reilly, Kevin van Zandvoort, C Julian Villabona-Arenas, Stefan Flasche, Rein M G J Houben, Hamish P Gibbs, Yang Liu, Samuel Clifford, Stéphane Hué, Alicia Rosello, Charlie Diamond, Sam Abbott, Quentin J Leclerc, Alicia Showering, Sophie R Meakin, Gwenan M Knight, Yung-Wai Desmond Chan, Sebastian Funk, Rosalind M Eggo, Thibaut Jombart, Nikos I Bosse, Christopher I Jarvis, Fiona Yueqian Sun, Megan Auzenbergs, Katherine E. Atkins, Rosanna C Barnard, Petra Klepac, Oliver Brady, Anna M Foss, Matthew Quaife, Georgia R Gore-Langton, Frank G Sandmann, James W Rudge, Simon R Procter, Jack Williams, Mark Jit, Arminder K Deol, Damien C Tully, David Simons, Rachel Lowe, Yalda Jafari, Nicholas G. Davies, Emily S Nightingale, Jon C Emery.
